# Explainable Machine Learning for Early Detection of Mild Cognitive Impairment, Fall Risk, and Frailty Using Sensor-Based Motor Function Data

**DOI:** 10.64898/2025.12.23.25342943

**Authors:** Sonia Akter, Trent M. Guess, Shraboni Sarker, Samuel A. Hockett, Andrew M. Kiselica, Jamie B. Hall, Praveen Rao

## Abstract

**Objective:** This study aimed to design and evaluate an explainable machine learning (ML) framework that integrates sensor-based motor assessments with demographic and clinical data to identify early indicators of mild cognitive impairment (MCI), fall risk, and frailty in older adults.

**Methods:** Eighty-three community-dwelling older adults (60 years or older) completed multimodal motor assessments using the Mizzou Point-of-Care Assessment System (MPASS) to capture synchronized gait, balance, and sit-to-stand performance. Sensor-derived motor features were combined with demographic and clinical variables to develop predictive models for MCI, frailty, and fall risk using XGBoost and Decision Tree algorithms. A unified multilabel framework was also developed using XGBoost, Decision Tree, and AdaBoost to predict all three outcomes. Model interpretability was evaluated using SHapley Additive exPlanations (SHAP).

**Results:** The ML model for MCI achieved the highest performance (94% accuracy, AUC = 0.88, F1 = 0.94), followed by fall risk (94% accuracy, AUC = 0.90) and frailty (82% accuracy, AUC = 0.77). Unified multilabel models showed moderate performance (67-73% accuracy), with XGBoost achieving the highest accuracy (73%), sensitivity, and F1 score, while the Decision Tree showed higher discrimination (AUC = 0.72). SHAP analyses identified stride length and time, center-of-pressure-based balance measures, and knee angular velocity during sit-to-stand as dominant predictors.

**Conclusions:** This work introduces a novel ML framework using multimodal sensor-based motor assessments to predict MCI, fall risk, and frailty individually and within a unified model. By combining explainable ML with motor-function data, the framework supports transparent early screening of multidomain cognitive and physical decline in aging.

## INTRODUCTION

Global population aging is contributing to increased public-health challenges linked to neurodegenerative and functional decline.^1^ Common geriatric syndromes, include mild cognitive impairment (MCI), frailty, and fall risk, represent interconnected domains of vulnerability that often precede and accelerate later cognitive and physical decline.^2–9^ First, MCI is an intermediate stage between normal aging and dementia, characterized by cognitive deterioration despite preserved independence, and associated with a substantially elevated risk of progression to dementia.^4,5^ Next, falls are a major source of injury and loss of independence among older adults, frequently leading to hospitalization, disability, and premature death.^7,8^ Finally, frailty is defined by reduced physiological reserve and multisystem dysregulation, increasing susceptibility to disability and death.^6,10^ These conditions are strongly interrelated: MCI predicts future falls; frailty and cognitive decline frequently co-occur; and recurrent falls exacerbate both physical and cognitive deterioration.^8,9^ Together, these conditions represent a multidomain vulnerability contributing to diminished quality of life and increased healthcare burden among older adults.^11,12^, underscoring the importance of early identification. Traditional screening approaches, including brief cognitive tests, gait-speed measurement, and the Timed Up-and-Go (TUG) test, may be insensitive to subtle early changes and difficult to scale for community-level use.^13–15^ Growing evidence suggests that motor-function alterations can emerge years before overt cognitive symptoms, positioning gait, balance, and sit-to-stand (STS) performance as early indicators of cognitive impairment, frailty, and fall risk.^16–19^ These findings highlight motor function as a promising digital biomarker for early detection of age-related cognitive and physical decline.

Recent advances in sensor technologies, including depth cameras, wearable inertial sensors, and force plates, enable high-resolution assessment of movement, balance, and reaction-time parameters across clinical and community settings.^20–22^ However, the high dimensionality and nonlinear structure of sensor-derived data necessitate advanced computational methods. Machine-learning (ML) approaches, particularly tree-based ensemble models such as Decision Trees^23^, AdaBoost^24^, and XGBoost^25^, have shown strong performance in modeling complex motor patterns and predicting geriatric outcomes.^2,3,33–38,25–32^

Despite these advances, many ML models function as “black boxes,” offering accurate predictions but limited transparency, reducing clinician trust and adoption in healthcare applications.^39,40^ Explainable ML methods address this gap by linking predictive performance with interpretable, physiologically meaningful insights.^18,27^, which is particularly critical for geriatric screening application.

### Objective

Given the need to improve early detection of geriatric clinical syndromes and the strong promise of sensor-based ML approaches to meet this need, the objective of this study was to design and evaluate an explainable ML framework that combines sensor-based motor function features with demographic and clinical data to identify early indicators of MCI, frailty, and fall risk in community-dwelling older adults.

## MATERIALS AND METHODS

### Participants

A total of 83 community-dwelling older adults aged 60+ years were enrolled in this study from the University of Missouri (MU) Health Psychology Clinic and surrounding mid-Missouri communities (MU IRB No. 2102007). Eligible individuals were required to walk independently without assistive devices and have no orthopedic or neurological conditions that could interfere with movement performance.

All participants completed a demographic and health history questionnaire along with the Montreal Cognitive Assessment (MoCA) to evaluate global cognitive function. Based on assessment outcomes, participants were independently labeled for each outcome of interest, with corresponding control groups defined for participants who did not meet outcome-specific criteria: assigned to different categories.

1. MCI: Defined as a MoCA score < 26 (Nasreddine et al., 2005)^41^ or a confirmed clinical diagnosis of MCI by a board-certified neuropsychologist. Participants who did not meet these criteria were labeled as cognitively unimpaired controls.
2. Fall risk: Defined as self-reported history of one or more falls within the previous 12 months, consistent with published prevalence benchmarks (25–30% of older adults) from national surveillance.^42^ Participants reporting no falls in the prior year were labeled as low fall-risk controls.
3. Frailty: Defined by using the Tilburg Frailty Indicator (TFI), with a score ≥ 5 indicating frailty.^43^ Participants with TFI scores below this threshold were labeled as non-frail controls.

Note that a participant may be assigned to zero, one, two, or all three categories.

### Data Collection

Motor function data were collected using the Mizzou Point-of-Care Assessment System (MPASS)^22,44–46^, a novel portable multimodal platform (Figure 1) designed to capture synchronized kinetic, kinematic, and cognitive performance measures. The system integrates a depth camera (30 Hz), a custom-built force plate, and an interactive interface board, enabling comprehensive assessment of gait, balance, sit-to-stand (STS), step-down, and reaction time tasks.

**Figure 1.**
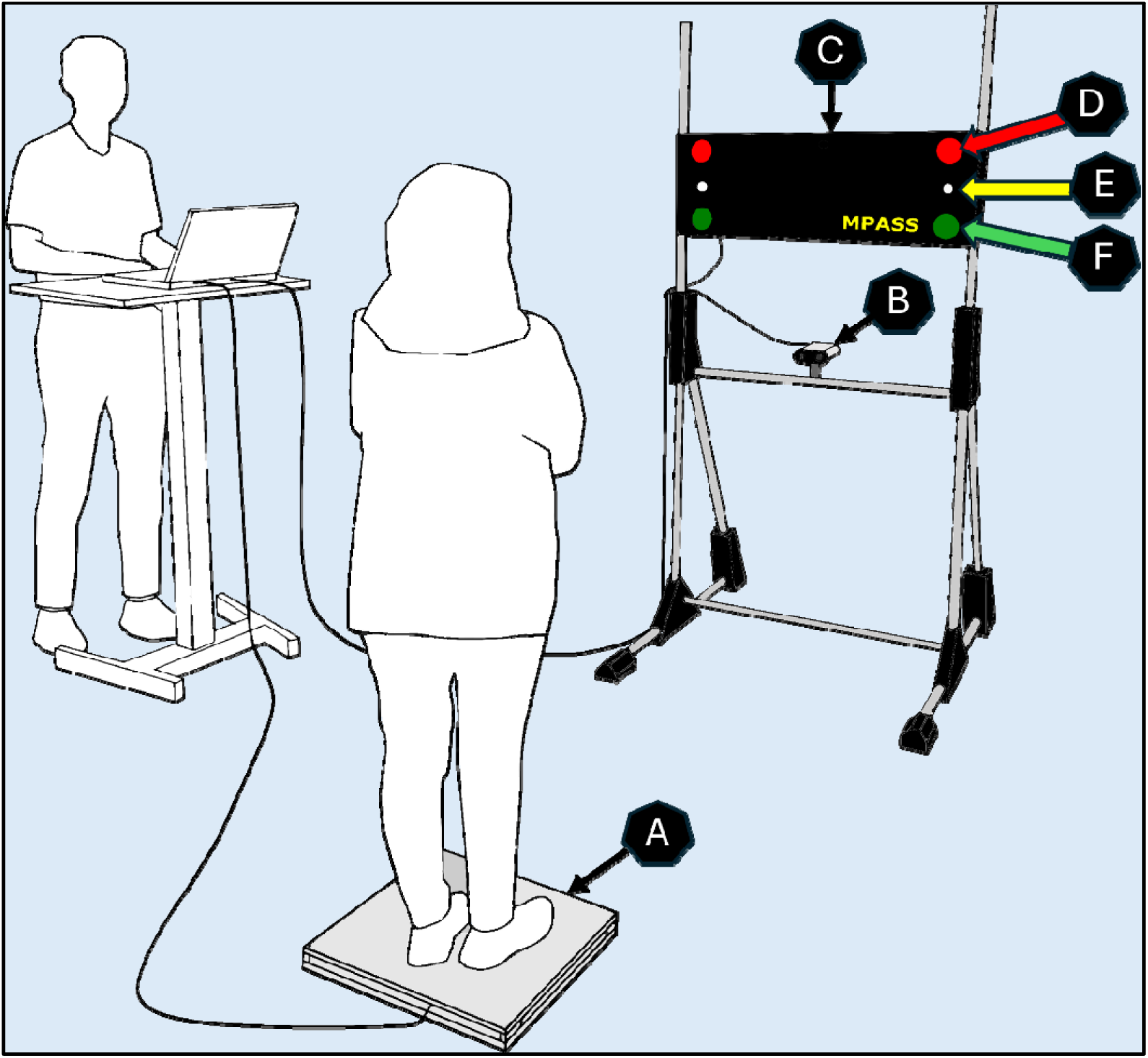
MPASS: Mizzou Point-of-care Assessment System. A - Custom Force Plate; B - Depth Camera; C – Interface Board; D – LED Red flashlight; E – Stop light; F – LED Green flashlight.

Each participant completed a standardized sequence of tasks representing multiple domains of physical function:

- Gait: Participants walked a 6-meter path three times at a standardized pace instructed by the examiner to ensure consistency across participants. The depth camera continuously tracked center-of-mass displacement and foot-placement trajectories to derive spatiotemporal gait parameters, including stride length, stride time, Step width, and Step length. ^44^
- STS: Participants rose from a 44 cm bench five consecutive times with arms crossed, assessing lower-limb strength, transition smoothness, and movement control. Transition smoothness was characterized using time-to-completion and average knee angular velocity during sit-to-stand transitions.^45^
- Static balance: Participants completed static balance trials while standing quietly on a force plate under single- and dual-task conditions. Balance performance was quantified using force-plate–derived center-of-pressure (CoP) sway area and velocity, including anteroposterior and mediolateral components, with additional balance features and definitions provided in the Supplementary Material.
- Step-Down: From a 21-cm platform, participants stepped down toward the depth camera three times per leg. Step-down performance was characterized using temporal measures for the leading and trailing limbs and mediolateral center-of-mass displacement during the task.
- Reaction-time: Using the MPASS interface board, participants responded to randomized LED flashes (red, green) by pressing the corresponding buttons, allowing quantification of reaction time and response accuracy.

To assess motor–cognitive interaction, gait, balance, and sit-to-stand tasks were repeated under a dual-task condition involving serial-seven subtraction, allowing evaluation of attentional demands and motor adaptability during divided-attention performance. All assessments were conducted under standardized conditions, with the depth camera positioned approximately 1 m above ground level and 2.5 m in front of the force plate to ensure consistent whole-body motion capture across participants.

#### Detailed definitions of all sensor-derived features are provided in the Supplementary Material

Demographic and clinical information were collected through standardized questionnaires administered during the study visit. Variables included age, sex, race/ethnicity, height, weight, and body mass index (BMI). Participants also reported family history of neurological and psychiatric conditions, including Alzheimer’s disease, dementia, Parkinson’s disease, stroke, cognitive impairment, schizophrenia, bipolar disorder, and alcoholism. These demographic and clinical variables were merged with sensor-derived motor features for subsequent analyses and model development.

#### Feature Engineering

Raw sensor signals collected from the MPASS platform were transformed into structured, task-specific features suitable for ML analysis. Feature extraction captured temporal, kinematic, and kinetic characteristics of motor performance across gait, sit-to-stand, static balance, step-down, and reaction-time tasks.

For each task and condition (single-task and dual-task), features were computed at the trial level and aggregated across repeated trials to obtain a single representative value per participant. Feature engineering emphasized clinically interpretable measures of movement timing, velocity, stability, symmetry, and motor control, while excluding raw sensor waveforms.

The final feature set used for model training to ensure consistency across participants and outcomes. A complete description of engineered features, grouped by task and condition, is provided in the Supplementary Materials (Supplementary Table S1-Table S5).

#### Data Preprocessing

Following data acquisition, all raw MPASS recordings were exported and processed in MATLAB (MathWorks, Natick, MA) to compute discrete motor and task-performance features for each assessment trial. These features represented multiple motor domains, including balance/postural stability, gait, STS performance, step-down, and reaction time. For each task and condition, repeated trials were averaged to obtain a single representative value per participant. MATLAB-derived feature files were then merged with demographic and clinical variables for analysis.

All features underwent systematic quality control to identify outliers, incomplete entries, and measurement anomalies. Missing values were imputed using the mean for normally distributed features and the median for skewed distributions. For participants unable to complete specific tasks, a sentinel value (-999) was assigned, together with a binary completion flag, were assigned to distinguish task non-completion from missing data. This approach is widely used in data mining and ML workflows, including *scikit-learn-based* pipelines, to preserve dataset completeness without exclusion bias.^47,48^ Categorical variables were one-hot encoded for ML compatibility.^49^

Three binary target variables were defined to represent each clinical outcome: for the MCI group, “Control” was encoded as 0 and “MCI” as 1; for fall risk, “No” (no falls in the last year) was encoded as 0 and “Yes” (one or more falls in the last year) as 1; and for frailty, “Non-frail” was encoded as 0 and “Frail” as 1. Two modeling strategies were evaluated: (1) Separate ML models to predict each outcome and (2) a unified multi-label model jointly predicting all three outcomes from shared features.

A total of 83 participants were included in the study. Fall-risk and frailty outcomes were available for all participants, whereas MCI classification required sufficient cognitive assessment data; four participants with insufficient cognitive assessment information were not excluded in MCI analysis. Thus, the MCI ML model included 79 participants (44 controls and 35 MCI). A random subset of 16 participants (8 controls and 8 MCI) was used as the hold-out test set (∼20%), and the remaining 63 participants (36 controls and 27 MCI) were used for model training (∼80%). To handle class imbalance, the training set was augmented using the Synthetic Minority Oversampling Technique (SMOTE).^50,51^

The fall-risk and frailty ML models, data from 83 participants were analyzed. The fall-risk model comprised of 50 non-fallers and 33 fallers; the frailty model included 66 non-frail and 17 frail participants. Both datasets were divided into 80% training and 20% testing, maintaining proportional class distributions. SMOTE was not applied, as analysis using the original unbalanced data yielded more stable and generalizable results.

The unified multi-label model included all 83 participants using stratified sampling, 80/20 splits preserving label co-occurrence. All data preprocessing and feature engineering were performed in Python (version 3.9.13) using Jupyter Notebook, with core libraries including pandas, NumPy, and *scikit-learn*.

#### Model Development and Validation

Next, we discuss how ML models were trained, tuned, and tested on the preprocessed data. We employed a combination of linear and ensemble-based classifiers to capture both linear and nonlinear relationships among multimodal motor, demographic, and clinical features. Logistic Regression (LR) served as the baseline model because of its simplicity, interpretability, and established role in clinical prediction modeling.^52^ Because motor-performance and frailty-related features often exhibit nonlinear dependencies and multicollinearity, tree-based and ensemble methods were implemented to improve predictive accuracy and generalizability.

The Decision Tree classifiers were selected for the rule-based structure, ability to handle mixed feature types, and interpretability in small, heterogeneous biomedical datasets.^28,33^ AdaBoost iteratively combines weak learners to enhance model stability, particularly under imbalance.^24,31,53^ The XGBoost algorithm was chosen for its computational efficiency, regularization, and strong performance in handling high-dimensional and nonlinear data.^25^ Together, these models provided complementary perspectives, balancing interpretability and predictive complexity across both single and unified classification frameworks.

Model training and evaluation were conducted for both separate ML models (MCI, fall risk, and frailty) and the unified multi-label model. For the separate models, LR classifiers were first trained for each outcome using identical data splits to establish a linear baseline; despite lower performance, LR was retrained as a benchmark to highlight gains from nonlinear methods. Single outcome models were tuned using GridSearchCV with stratified 10-fold cross-validation, while the unified multilabel model was optimized using RandomizedSearchCV with stratified 3-fold cross-validation within a MultiOutputClassifier framework. Hyperparameter configurations are summarized in Supplementary Table S6.

The unified multilabel model jointly predicted MCI, fall risk, and frailty using shared features. XGBoost, Decision Tree, and AdaBoost were evaluated within the MultiOutputClassifier framework, with XGBoost selected as the final unified classifier based on cross-validation performance. After optimization, the model was retrained on the full training set and evaluated on the test set.

Model explainability was assessed using SHapley Additive exPlanations (SHAP), which provides global and instance-level interpretations of a complex model. ^3,26,27^ SHAP analyses were applied to the best-performing separate and unified models to quantify feature contributions and identify shared and outcome-specific predictors associated.

#### Performance Evaluation

Model performance was assessed using metrics such as accuracy, area under the receiver operating characteristic curve (AUC–ROC), precision, sensitivity, specificity, and F1-score. For the unified model, each metric was calculated as a weighted average across the three outcomes to account for class imbalance and ensure balanced evaluation across outcomes. Predicted probabilities were classified using a fixed threshold of 0.5, where values ≥ 0.5 were labeled as positive and < 0.5 as negative for each outcome.

To further evaluate model behavior and robustness, post-model analyses were performed to examine feature relevance, statistical significance, and generalizability. Feature-importance values were extracted from the trained models to identify the most influential motor, demographic, and clinical predictors associated with each outcome. Statistical comparisons were then conducted to test significant differences among the top-ranked features contributing to classification.

#### ML Workflow Overview

Figure 2 summarizes the overall ML workflow used in this study, outlining the sequential steps of data collection, data preprocessing, feature standardization, model selection, model training with cross-validation and hyperparameter tuning, and post hoc model interpretation using SHAP.

**Figure 2.**
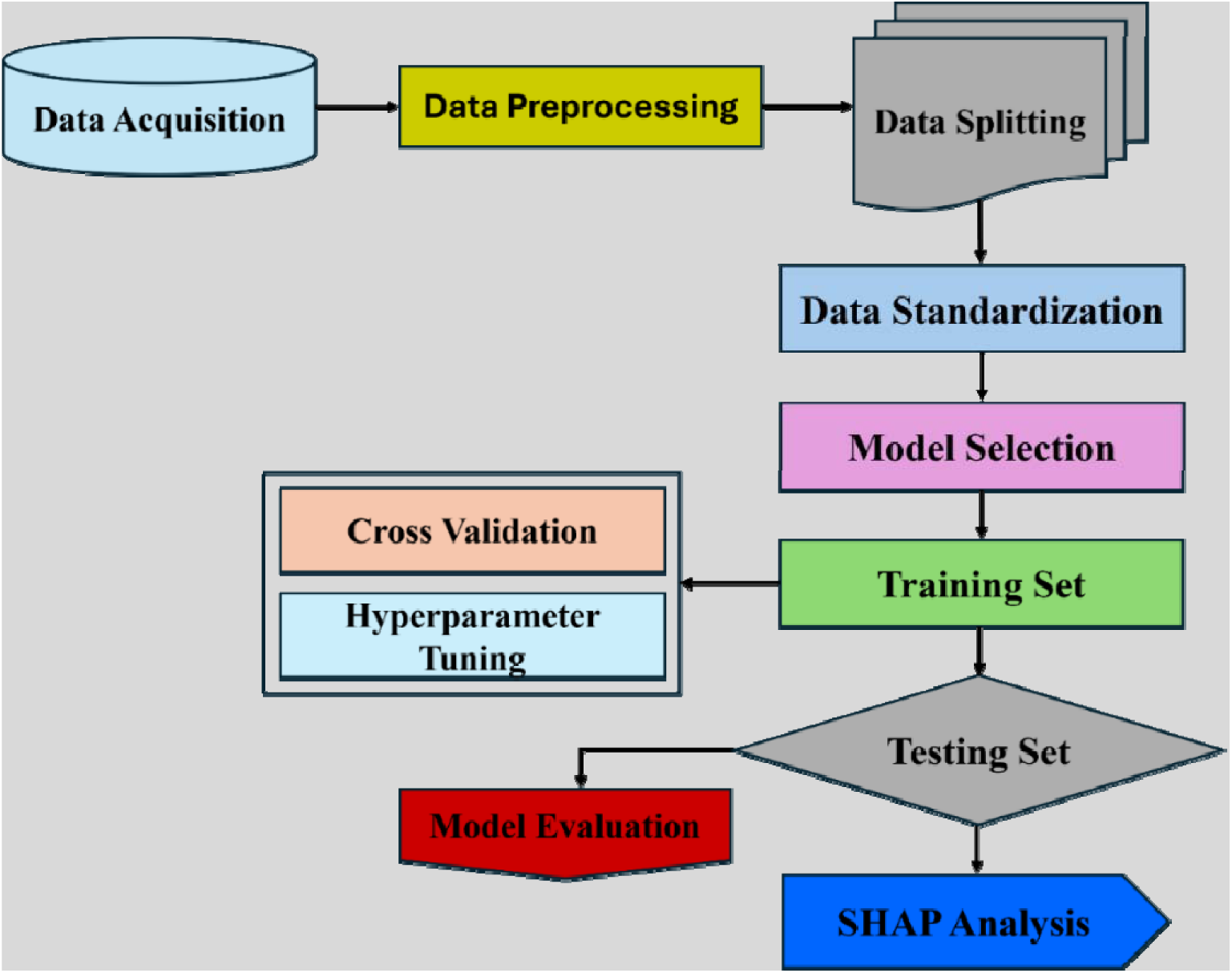
Analytical workflow of the machine learning (ML) model study.

## RESULTS

### Participants Characteristics

A total of 83 community-dwelling older adults (≥60 years) were included (mean age 70.7 ± 4.7 years; ∼60% female). The cohort was predominantly White (96.4%). Mean height, weight, and BMI were 1.69 ± 0.10 m, 79.5 ± 16.3 kg, and 28.0 ± 5.2 kg/m², respectively (Table 1).

**Table 1.** Descriptive statistics by participant groups (MCI, Fall Risk, and Frailty)*.

| **MCI** | **Overall (n = 79)** | **Control (n = 44)** | **MCI (n = 35)** | **p-value** |
| --- | --- | --- | --- | --- |
| **Characteristics** |  |  |  |  |
| Age (years) | 70.86 ± 4.68 | 69.93 ± 4.08 | 72.03 ± 5.17 | 0.0473 |
| Sex (female) | 48 (60.8%) | 30 (68.2%) | 18 (51.4%) | 0.166 |
| **Race** |  |  |  | 0.358 |
| White | 77 (97.5%) | 43 (97.7%) | 34 (97.1%) | N/A |
| Black or AA | 1 (1.3%) | 0 | 1 (2.9%) | N/A |
| Other | 1 (1.3%) | 1 (2.3%) | 0 | N/A |
| Height (m) | 1.68 ± 0.10 | 1.69 ± 0.09 | 1.68 ± 0.11 | 0.773 |
| Weight (kg) | 79.5 ± 16.6 | 75.6 ± 13.9 | 84.4 ± 18.5 | 0.0178 |
| BMI (kg/m<sup>2</sup>) | 28.0 ± 5.2 | 26.5 ± 4.6 | 29.8 ± 5.4 | 0.00462 |
| **Fall Risk** | **Overall (n = 83)** | **Non-fall (n = 50)** | **Fall (n = 33)** | **p-value** |
| **Characteristics** |  |  |  |  |
| Age (years) | 70.71 ± 4.70 | 71.22 ± 5.12 | 69.94 ± 3.93 | 0.227 |
| Sex (female) | 50 (60.2%) | 27 (54.0%) | 23 (69.7%) | 0.175 |
| **Race** |  |  |  | 0.417 |
| White | 80 (96.4%) | 48 (96.0%) | 32 (97.0%) | N/A |
| Black or AA | 1 (1.2%) | 1 (2.0%) | 0 | N/A |
| Other | 1 (1.2%) | 1 (2.0%) | 0 | N/A |
| Height (m) | 1.69 ± 0.10 | 1.69 ± 0.10 | 1.67 ± 0.10 | 0.345 |
| Weight (kg) | 79.5 ± 16.3 | 81.6 ± 15.2 | 76.2 ± 17.5 | 0.139 |
| BMI (kg/m<sup>2</sup>) | 28.0 ± 5.2 | 28.6 ± 5.5 | 27.0 ± 4.6 | 0.183 |
| **Frailty** | **Overall (n = 83)** | **Non-frail (n = 61)** | **Frail (n = 22)** | **p-value** |
| **Characteristics** |  |  |  |  |
| Age (years) | 70.71 ± 4.70 | 70.67 ± 4.15 | 70.82 ± 6.08 | 0.901 |
| Sex (female) | 50 (60.2%) | 35 (57.4%) | 15 (68.2%) | 0.451 |
| **Race** |  |  |  | 0.321 |
| White | 80 (96.4%) | 59 (96.7%) | 21 (95.5%) | N/A |
| Black or AA | 1 (1.2%) | 1 (1.6%) | 0 | N/A |
| Asian | 1 (1.2%) | 1 (1.6%) | 0 | N/A |
| Other | 1 (1.2%) | 0 | 1 (4.5%) | N/A |
| Height (m) | 1.69 ± 0.10 | 1.70 ± 0.09 | 1.66 ± 0.12 | 0.122 |
| Weight (kg) | 79.5 ± 16.3 | 78.9 ± 15.6 | 81.2 ± 18.2 | 0.571 |
| BMI (kg/m<sup>2</sup>) | 28.0 ± 5.2 | 27.4 ± 5.0 | 29.5 ± 5.4 | 0.109 |

\*Values are presented as n (%) for categorical variables and mean ± SD for continuous variables. Group comparisons were conducted using Pearson's Chi-squared or Fisher's exact test for categorical data and one-way ANOVA for continuous variables. AA = African American.

For MCI prediction (n = 79; 44 controls, 35 MCI), participants with MCI were slightly older and had higher weight and BMI than controls (p < 0.05), while no differences were observed for sex, race, or height. For fall risk (n = 83; 50 non-fallers, 33 fallers) and frailty (n = 83; 61 non-frail, 22 frail), no significant group differences were observed in demographic or anthropometric variables. Overall, age and body composition primarily distinguished groups, providing context for subsequent ML analyses.

### Performance of Separate ML Models

The performance of separate ML models for predicting MCI, fall risk, and frailty is summarized in Table 2 and Figure 3. Model performance was evaluated using accuracy, AUC–ROC, precision, sensitivity, and F1-score.

**Table 2.**
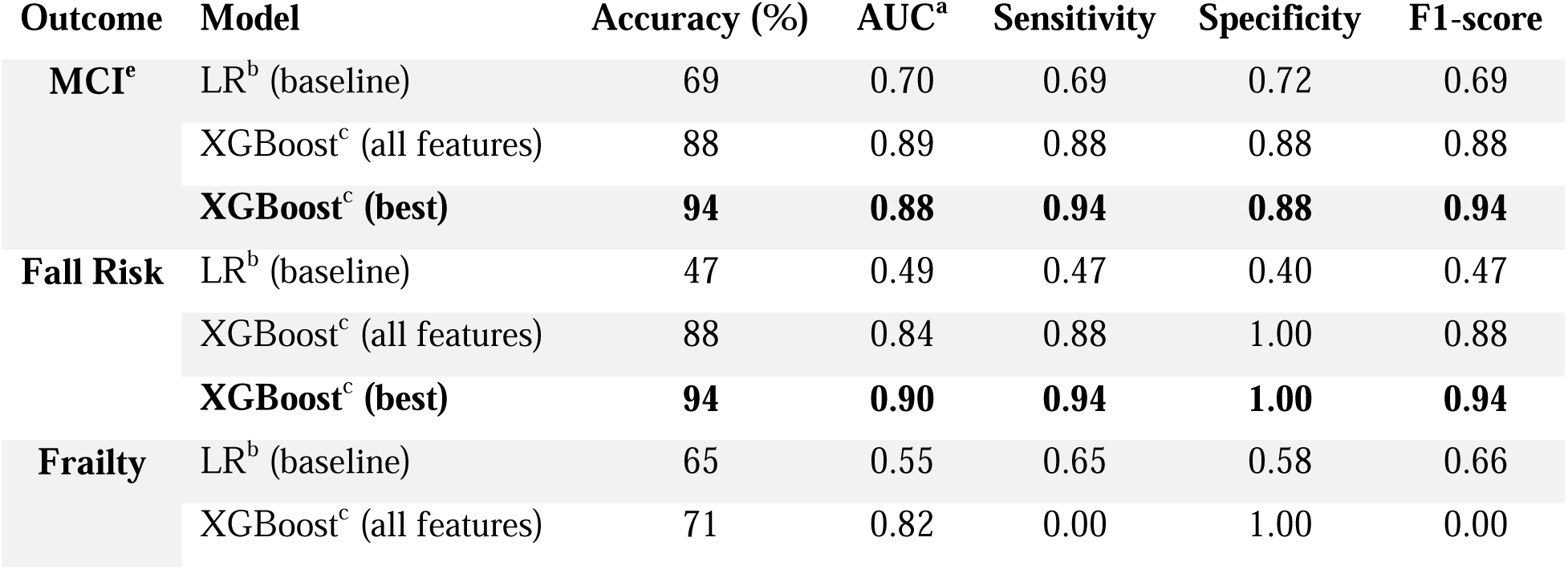
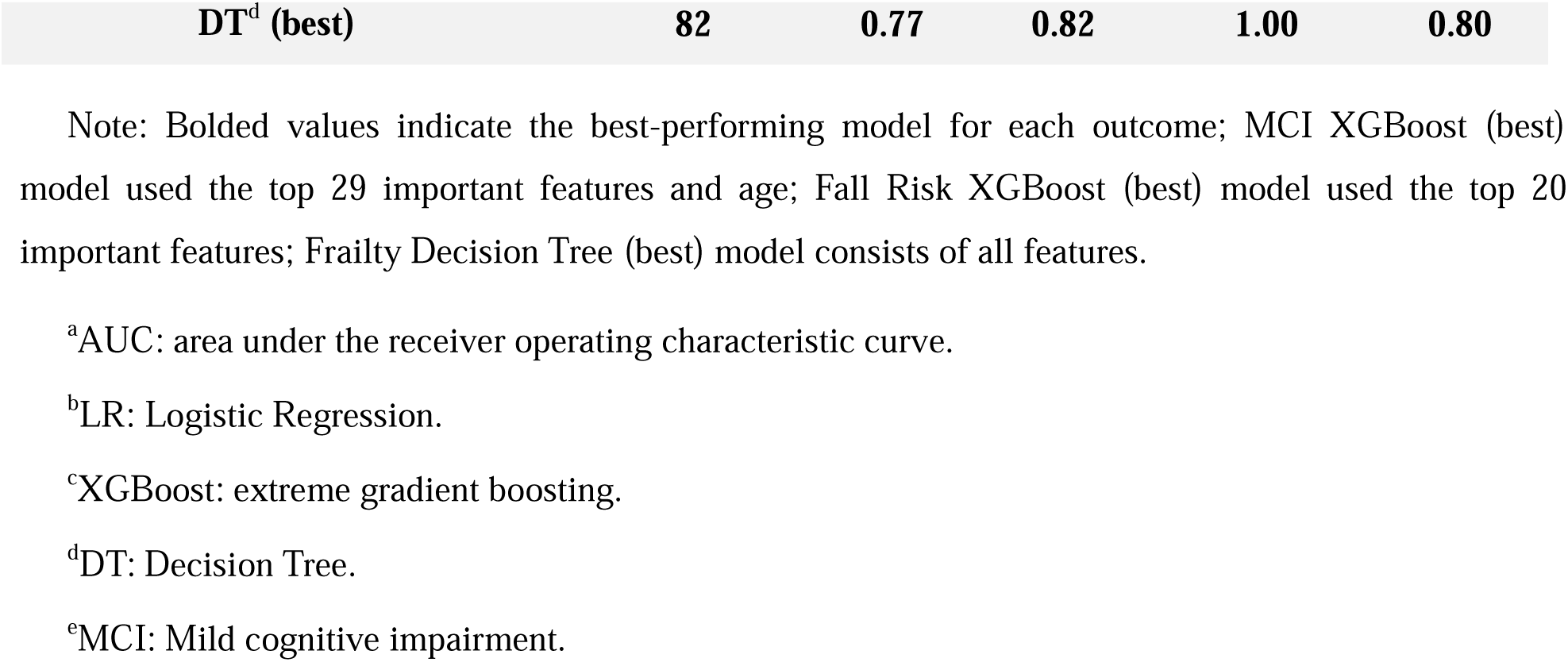
Performance of MCI, fall risk, and frailty predictive models (baseline, all features, and best models).

**Figure 3.**
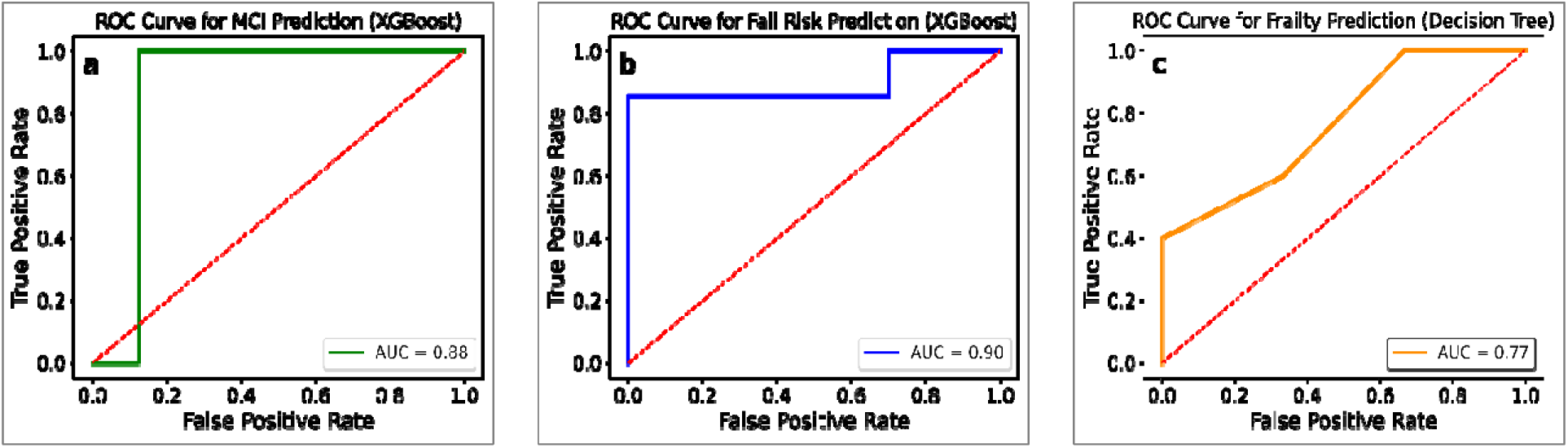
Receiver operating characteristic (ROC) curves for the best-performing machine-learning models predicting (a) mild cognitive impairment (MCI) using XGBoost, (b) fall risk using XGBoost, and (c) frailty using a Decision Tree. The dashed diagonal line indicates no-discrimination performance.

### MCI Prediction

The baseline logistic regression model, trained on all features, showed moderate discriminative ability, indicating limited linear separability between participants with MCI and cognitively unimpaired controls (Table 2).

When trained on the same feature set, XGBoost substantially improved classification performance, highlighting the importance of capturing nonlinear relationships among multimodal motor features. Building on this result, feature importance analysis based on cumulative gain was applied to identify the most informative predictors. A reduced feature set comprising the top 29 features (≥90% cumulative importance) along with age was then used to construct the final model (Supplementary Figure S7). This feature-selected XGBoost model achieved the strongest overall performance (Table 2; Figure 3a), with improvements confirmed by permutation testing (p = 0.002; Supplementary Figure S8).

To examine generalizability, the XGBoost (all features) classifier was evaluated across five independent test sets. Performance remained consistent (mean ± SD: accuracy 89 (%) ± 0.02, AUC = 0.93 ± 0.04), demonstrating that the model robustly captured reproducible motor-cognitive patterns rather than overfitting to a specific data split. See Table 3.

**Table 3.** Model generalizability across five randomly selected test sets (MCI model).

| Metric | Test 1 | Test 2 | Test 3 | Test 4 | Test 5 | Mean ± SD |
| --- | --- | --- | --- | --- | --- | --- |
| Accuracy (%) | 88 | 94 | 88 | 88 | 88 | $89 \pm 0.02$ |
| AUC | 0.95 | 1.00 | 0.92 | 0.86 | 0.94 | $0.93 \pm 0.04$ |
| Sensitivity | 0.88 | 0.94 | 0.88 | 0.88 | 0.88 | $0.89 \pm 0.02$ |
| Specificity | 0.75 | 1.00 | 0.88 | 0.75 | 0.88 | $0.85 \pm 0.09$ |
| Precision | 0.90 | 0.94 | 0.88 | 0.90 | 0.88 | $0.90 \pm 0.02$ |
| F1- score | 0.88 | 0.94 | 0.88 | 0.88 | 0.88 | $0.89 \pm 0.02$ |

### Fall Risk Prediction

The baseline LR model exhibited limited discriminative ability for classifying fallers vs. non-fallers (accuracy 47%, AUC = 0.49, F1 = 0.47; see Table 2), suggesting that the relationship between motor features and fall status was nonlinear. Consistent with this observation, the confusion matrix (Supplementary Figure S9) showed frequent misclassifications of fallers, further highlighting the limitations of linear modeling for this outcome.

In contrast, the XGBoost model trained using all features substantially improved predictive performance (accuracy 88%, AUC = 0.84, F1 = 0.88; Table 2), reflecting enhanced sensitivity and specificity relative to the baseline model. TO further improve model performance, feature importance analysis was conducted to identify the most informative predictors. As shown in Supplementary Figure S10, model performance varied with the number of selected features, with peak performance achieved using a reduced subset of top-ranked features.

The final XGBoost model, retrained using these top-ranked features, achieved the strongest performance, with an accuracy of 94%, an AUC of 0.90, and an F1 score of 0.94 (Table 2, Figure 3b). A permutation test confirmed that the observed AUC significantly exceeded chance performance (p < 0.01; Supplementary Figure S11), supporting the robustness and statistical significance of the final fall risk classifier.

### Frailty Prediction

For the frailty classification task, the baseline LR model demonstrated limited discriminative performance (accuracy = 65%, AUC = 0.55, F1 = 0.66; Table 2), suggesting that linear frailty-related patterns were not well captured by linear decision boundaries.

When trained using all features, the XGBoost model achieved a higher AUC (0.82; Table 2); However, it failed to correctly identify frail participants, resulting in zero sensitivity and an F-1 score of 0.00. Given this imbalance and lack of clinically meaningful detection of frailty cases, the XGBoost model was not considered suitable for this outcome despite its higher overall AUC.

A single DT classifier provided a more balanced and reliable performance, achieving an accuracy of 82%, an AUC of 0.77, a sensitivity of 0.82, and an F-1-score of 0.80 (Table 2; Figure 3c). Feature importance analysis conducted for the frailty task (Supplementary Figure S12) did not yield performance improvements when reduced feature subsets were used. Consequently, the final DT model was trained using the full feature set and selected as the best-performing and most stable classifier for frailty prediction.

### SHAP-Based Model Interpretability

Model interpretability was evaluated using SHAP to examine how individual features contributed to predictions of MCI, fall risk, and frailty. As shown in Figure 4, SHAP summary plots display participant-level feature effects, while bar plots summarize global feature importance based on mean absolute SHAP values.

**Figure 4.**
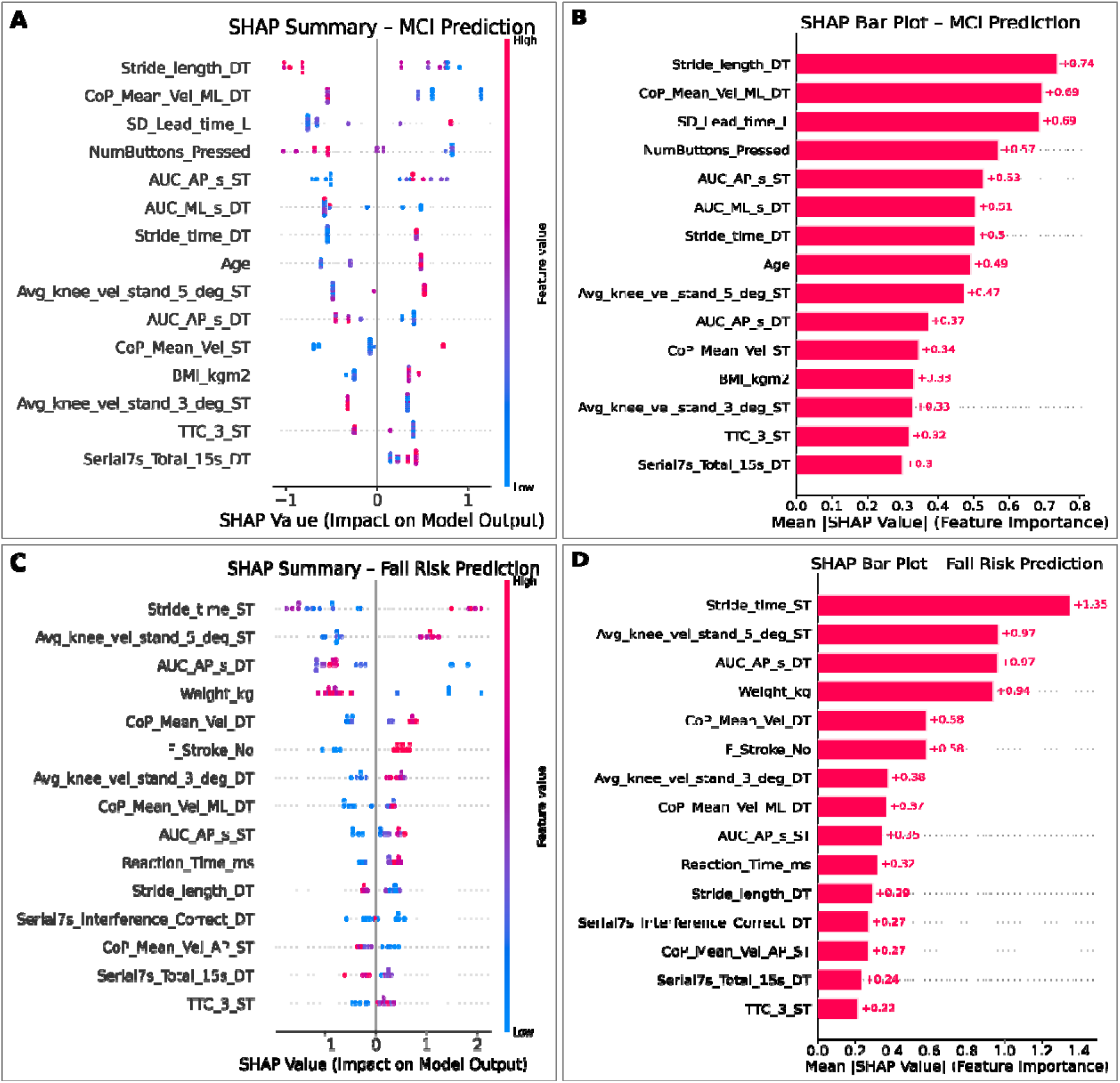
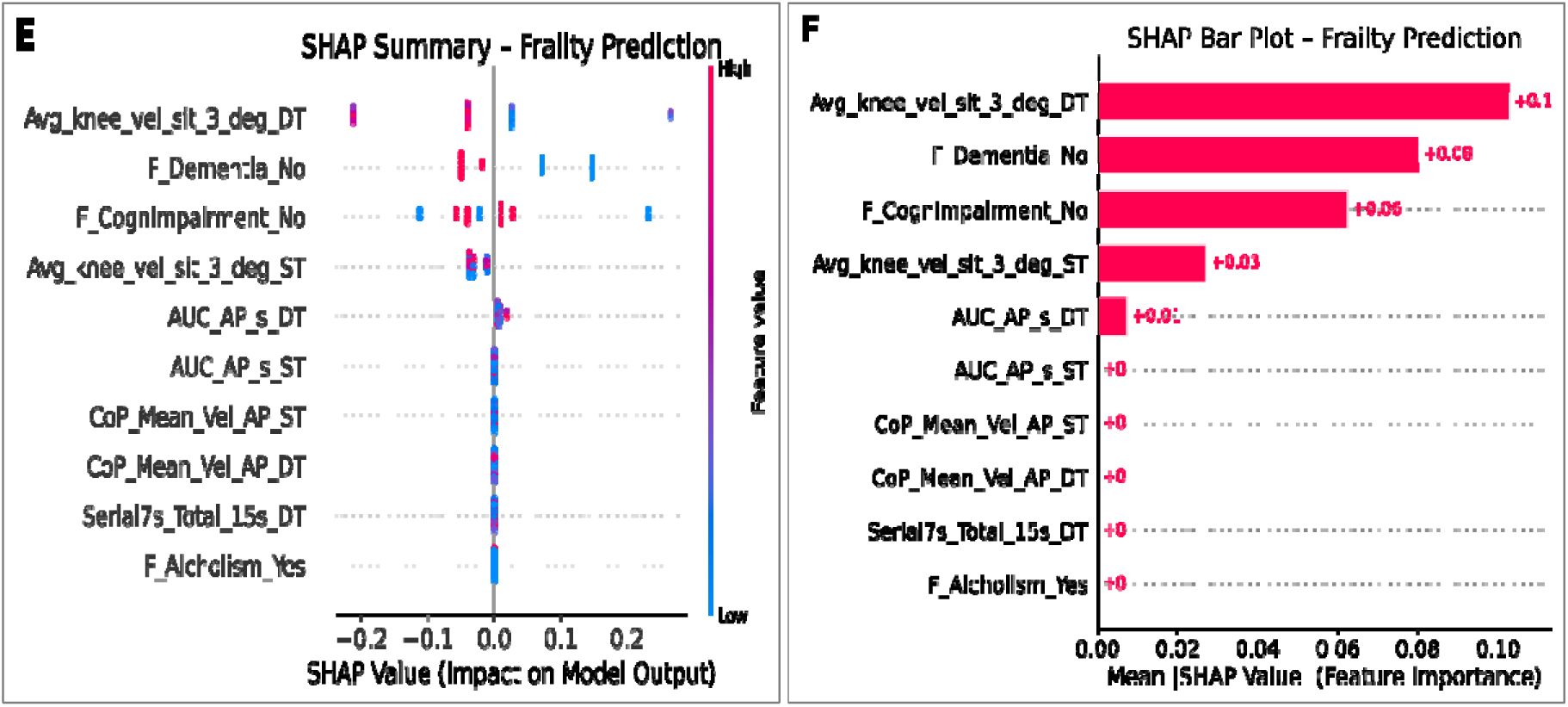
SHapley Additive exPlanations (SHAP) interpretation of the best-performing outcome-specific models. SHAP summary plots (left) and mean absolute SHAP value bar plots (right) are shown for prediction of (A–B) MCI, (C–D) fall risk, and (E–F) frailty. Across outcomes, demographic, and sensor-derived gait, balance, and sit-to-stand features consistently contributed to model predictions. Note: In the SHAP summary plots, each point represents a participant, with color indicating relative feature value (blue = low; red = high). Horizontal position reflects the magnitude and direction of feature contribution; positive SHAP values indicate increased predicted risk, and negative values indicate reduced risk.

For MCI prediction, the largest SHAP contributions arose from dual-task stride length, center-of-pressure mean velocity, particularly in the mediolateral direction, step-down timing of the leading limb, reaction-related button-press behavior, balance stability metrics (AUC), stride time, age, and knee angular velocity during STS. Higher stride length values were associated with negative SHAP values, shifting predictions toward the non-MCI class, whereas lower center-of-pressure velocity values shifted predictions toward MCI. For fall-risk prediction, stride time, knee angular velocity, balance stability metrics, body weight, center-of-pressure velocity, reaction time, and Serial 7s dual-task interference ranked highest in importance. For frailty prediction, knee angular velocity during STS emerged as the dominant contributor, followed by family history indicators of dementia or cognitive impairment. Overall, SHAP analyses highlighted both shared and outcome-specific motor, balance, cognitive, and demographic features driving model predictions.

### Unified ML Model Performance

After establishing optimized performance for outcome-specific models, we next evaluated whether MCI, fall risk, and frailty could be predicted jointly using a single machine-learning framework trained on shared sensor-derived motor features. Figure 5 illustrates this unified multi-label modeling approach, in which a common set of preprocessed features is used to generate simultaneous predictions for all three outcomes within a single pipeline.

**Figure 5.**
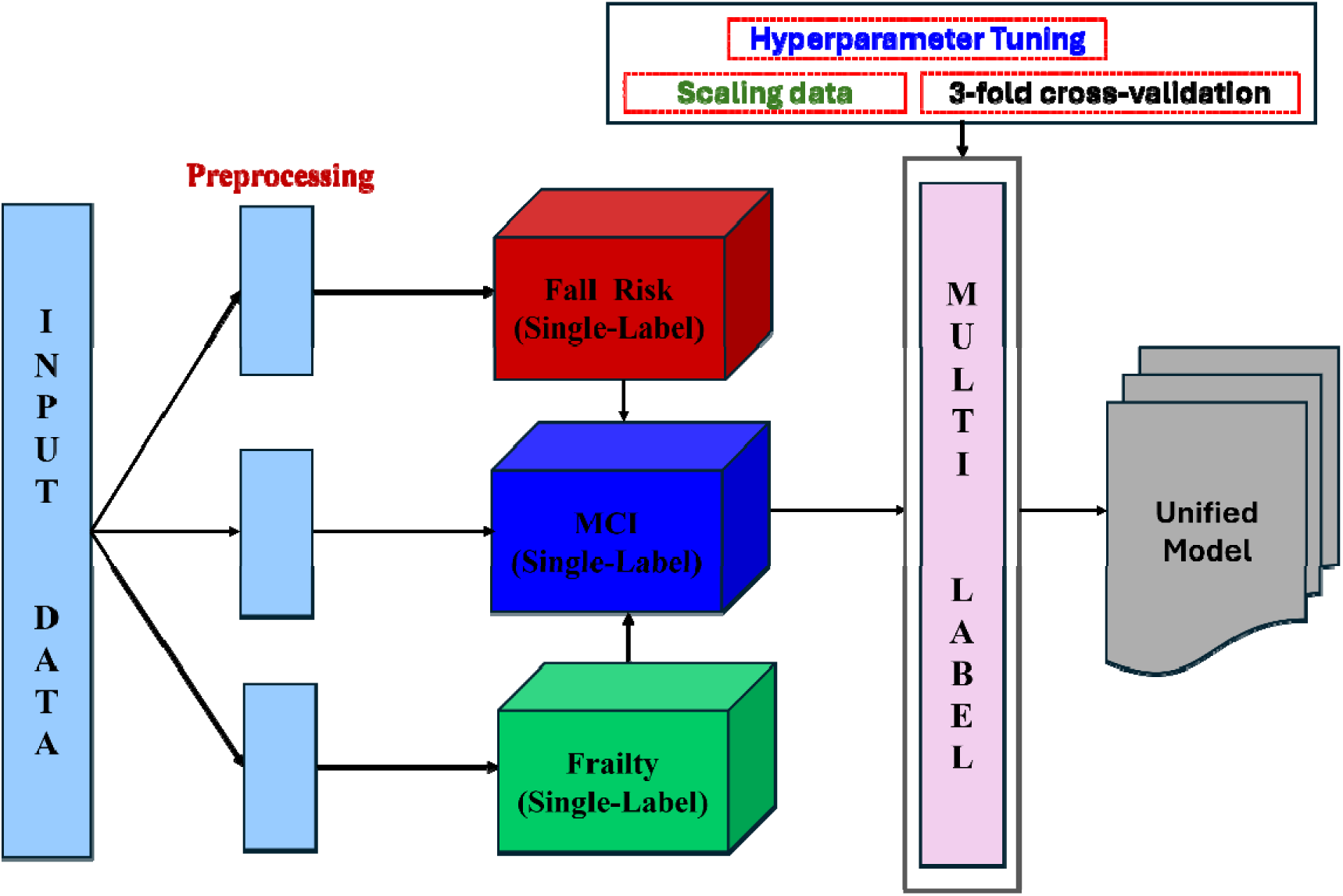
Unified multilabel model integrating MCI, Fall Risk, and Frailty into a single framework.

Unified multi-label models based on XGBoost, Decision Tree, and AdaBoost were trained and evaluated using the stratified train–test split described in Methods (Section 2.4). As summarized in Table 4, the unified models achieved moderate average weighted accuracy across outcomes (67–73%), with overall performance lower than that observed for the corresponding outcome-specific models. Across classifiers, the Decision Tree achieved the highest average weighted AUC, XGBoost demonstrated the highest weighted sensitivity, specificity, and F1 score, and AdaBoost showed comparable specificity with lower sensitivity.

**Table 4.**
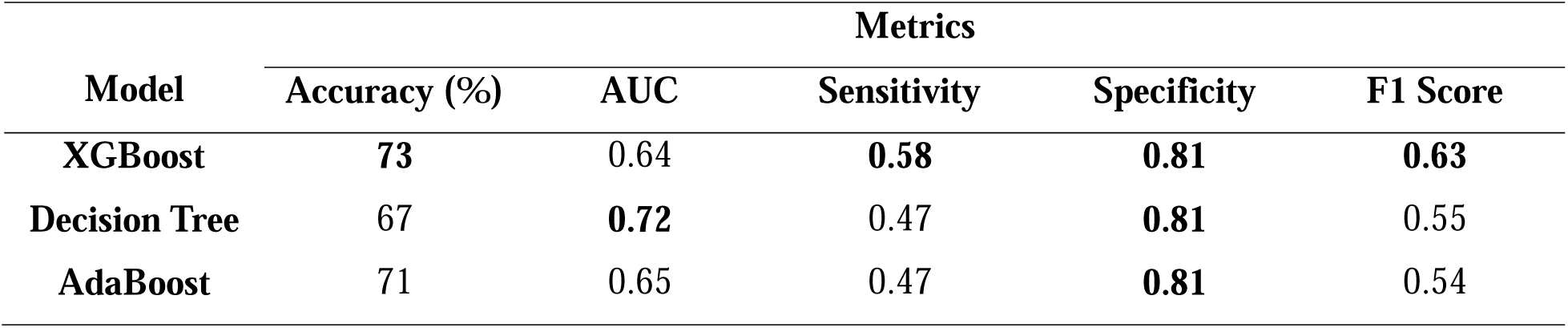
Average Weighted Performance (Across MCI, Fall Risk, and Frailty) of the Unified Multilabel Model.

Outcome-specific performance metrics underlying these averaged results are provided in Supplementary Table S13, and corresponding confusion matrices for the unified XGBoost and Decision Tree models are shown in Supplementary Figure S14.

### Model Explainability and SHAP-Based Interpretation

SHAP summary plots for the unified multi-label XGBoost and Decision Tree models are shown in Figure 6. In both models, features are ranked by mean absolute SHAP value, with point color indicating relative feature magnitude and horizontal position reflecting the direction and strength of contribution to predicted risk.

**Figure 6.**
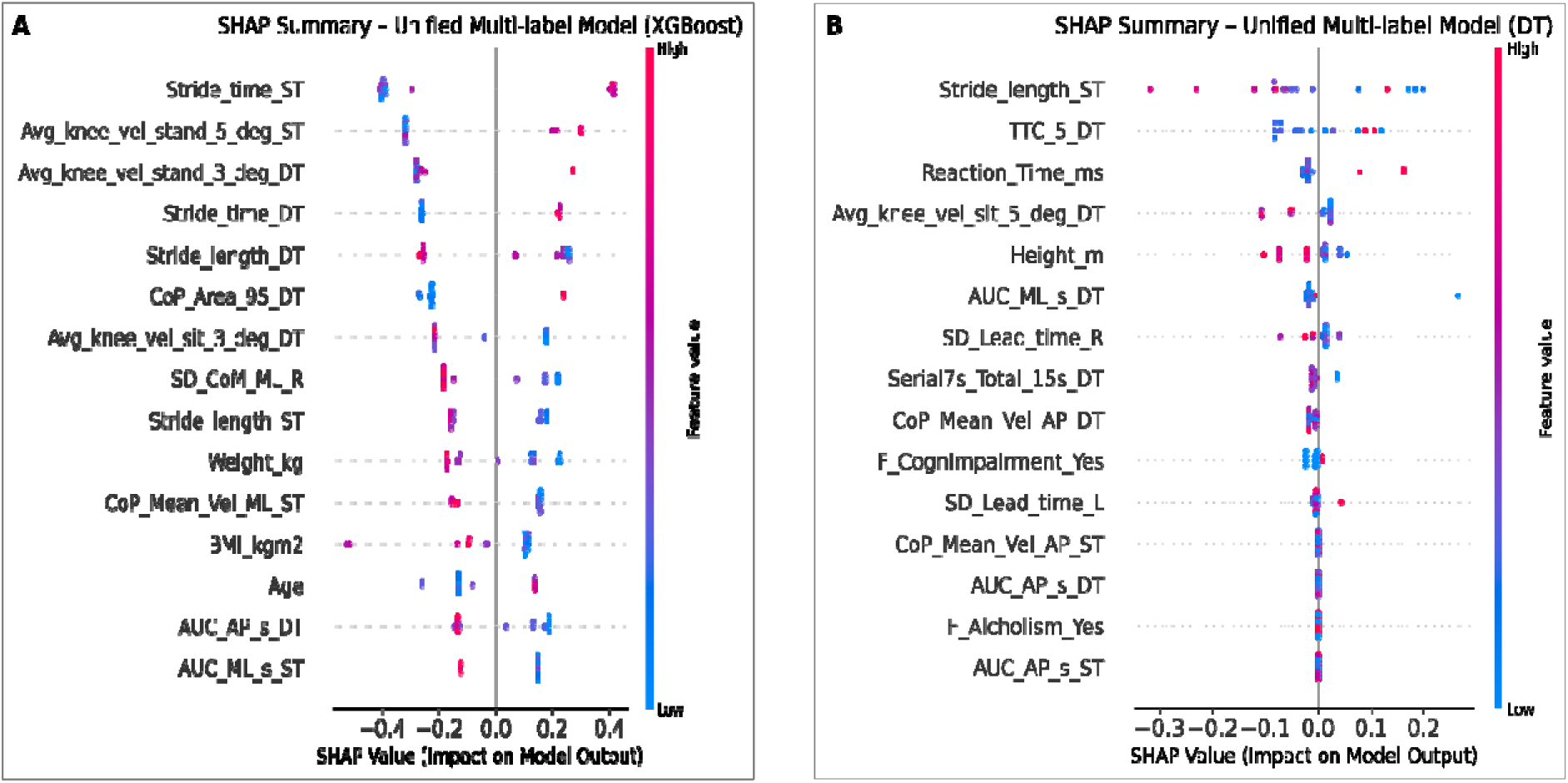
SHAP summary plots for unified multi-label models: (A) XGBoost model and (B) Decision Tree model.

Both unified models exhibited highly similar importance patterns, with overlapping top-ranked features and comparable directions of effect, consistent with their similar discriminative performance. The most influential features included gait measures (stride time and stride length), balance-related metrics (center-of-pressure area and velocity, AUC-based stability measures), STS kinematics (knee angular velocity), step-down timing features, and time-to-contact (TTC) measures. Demographic and anthropometric variables (age, height, weight, BMI), reaction time, and Serial 7s dual-task performance also contributed to the unified predictions.

Overall, longer stride time, greater balance instability, slower knee angular velocity, and delayed step-down or TTC measures were generally associated with increased predicted risk, whereas faster STS knee motion contributed negatively. These findings demonstrate that the unified model captures shared gait, balance, cognitive, and functional motor features that jointly influence the prediction of MCI, fall risk, and frailty.

## DISCUSSION

To our knowledge, this study is the first to jointly investigate predictive models for MCI, fall risk, and frailty using ML by training on sensor-based motor assessments. Across outcomes, nonlinear ensemble models outperformed LR, underscoring the nonlinear and interactive nature of motor and cognitive relation to disorder of aging. The MCI model achieved the strongest and most reproducible performance (94% accuracy), while thefall-risk model improved significantly after feature refinement (94% accuracy). Frailty model achieved robust performance (82% accuracy) using an optimized Decision Tree. The unified model preserved balanced discrimination while reducing pipeline redundancy, indicating that shared gait, balance, and dual-task features capture generalizable vulnerability across geriatric syndromes. For MCI, model-identified features aligned with prior evidence linking dual-task gait disruptions, shorter stride length, prolonged and irregular stride time, and increased variability to early cognitive impairment, likely reflecting competition for attentional and executive resources.^54^ Fall-risk predictions were driven primarily by postural sway metrics, consistent with established associations between impaired balance control and incident falls.^55^ Frailty prediction emphasized sit-to-stand performance, particularly reduced knee angular velocity, reflecting diminished lower-limb strength and functional reserve.^56^ Together, these findings indicate that disruptions in gait control, balance stability, and lower-limb power represent shared physiological mechanisms underlying multidomain vulnerability in aging.

Model interpretability was enhanced through SHAP analysis, which provided both global and participant-level explanations for the unified XGBoost model.^18^ SHAP revealed stable and physiologically coherent feature patterns across outcomes, strengthening transparency and clinical trust. These interpretable outputs may support actionable screening, longitudinal monitoring, and individualized feedback in older adults.

Several limitations should be noted. The study cohort was predominantly White, limiting generalizability and highlighting the need for validation in more diverse populations. Models relied on sensor-derived motor features and demographics; integrating complementary data sources such as electronic health records (EHRs) may further improve discrimination and calibration. Prior work has demonstrated that EHR-based phenotyping and longitudinal clinical data can enhance early detection and risk stratification for cognitive and functional decline.^57,58^

Future work will focus on prospective, longitudinal validation, expansion to larger and more diverse cohorts, and integration of multimodal data sources, including EHRs and cognitive assessments, to enhance robustness and clinical utility. Evaluating workflow feasibility will be essential for scalable deployment.

## CONCLUSIONS

In this study, we present an explainable ML framework for predicting MCI, fall risk, and frailty using sensor-derived motor assessments. By developing and evaluating separate outcome-specific models, we demonstrate that nonlinear ML approaches substantially outperform traditional LR, underscoring the importance of capturing complex motor–cognitive relationships in aging. Both the MCI and fall-risk models achieved strong performance, while the frailty model showed robust and clinically meaningful discrimination using task-specific motor features. A key contribution of this work is the systematic integration of multimodal motor tasks with SHAP-based interpretability to identify physiologically meaningful predictors, including gait dynamics and balance instability for MCI, postural stability and functional motor control for fall risk, and STS knee angular velocity for frailty. These interpretable patterns align with established clinical mechanisms and illustrate how transparent ML models can support early risk identification and informed screening in aging populations.

## Data Availability

The data used in this study are not publicly available at this time due to ethical and privacy considerations. Data may be made available from the corresponding author upon reasonable request once the work is published in peer reviewed journal, and subject to institutional approval.

## AUTHOR CONTRIBUTIONS

The research was designed by PR, TG, and SA.

Data collection was done by SA, SS, SH, JH, and TG.

The initial manuscript was drafted by SA; coding, data preprocessing, and data analysis were done by SA.

PR, TG, and AK edited the initial manuscript draft and provided feedback.

All authors reviewed and provided feedback for the final manuscript draft.

## SUPPLEMENTARY MATERIAL

Supplementary materials are available at the journal’s website.

## REFERENCES

1. Prince M, Bryce R, Albanese E, Wimo A, Ribeiro W, Ferri CP. The global prevalence of dementia: A systematic review and metaanalysis. Alzheimer’s Dement. 2013;9(1):63–75.e2. doi:10.1016/j.jalz.2012.11.007

2. Leghissa M, Carrera Á, Iglesias CA. Machine learning approaches for frailty detection, prediction and classification in elderly people: A systematic review. Int J Med Inform. 2023;178(July):105172. doi:10.1016/j.ijmedinf.2023.105172

3. Liang HW, Ameri R, Band S, et al. Fall risk classification with posturographic parameters in community-dwelling older adults: a machine learning and explainable artificial intelligence approach. J Neuroeng Rehabil. 2024;21(1):1–17. doi:10.1186/s12984-024-01310-3

4. Petersen RC, Doody R, Kurz A, et al. Current concepts in mild cognitive impairment. Arch Neurol. 2001;58(12):1985–1992. doi:10.1001/archneur.58.12.1985

5. Petersen RC. Mild cognitive impairment as a diagnostic entity. J Intern Med. 2004;256(3):183–194. doi:10.1111/j.1365-2796.2004.01388.x

6. Fried LP, Tangen CM, Walston J, et al. Frailty in older adults: Evidence for a phenotype. Journals Gerontol - Ser A Biol Sci Med Sci. 2001;56(3):146–157. doi:10.1093/gerona/56.3.m146

7. Ambrose AF, Paul G, Hausdorff JM. Risk factors for falls among older adults: A review of the literature. Maturitas. 2013;75(1):51–61. doi:10.1016/j.maturitas.2013.02.009

8. Verghese J, Wang C, Lipton RB, Holtzer R. Motoric cognitive risk syndrome and the risk of dementia. Journals Gerontol - Ser A Biol Sci Med Sci. 2013;68(4):412–418. doi:10.1093/gerona/gls191

9. Skonetzki S, Lüders F, Engelbertz C, et al. Aging and Outcome in Patients With Peripheral Artery Disease and Critical Limb Ischemia. J Am Med Dir Assoc. 2016;17(10):927–932. doi:10.1016/j.jamda.2016.06.004

10. Clegg A, Young J, Iliffe S, Rikkert MO, Rockwood K. Frailty in elderly people. Lancet. 2013;381(9868):752–762. doi:10.1016/S0140-6736(12)62167-9

11. 2024 Alzheimer’s disease facts and figures. Alzheimer’s Dement. 2024;20(5):3708–3821. doi:10.1002/alz.13809

12. Wimo A, Guerchet M, Ali GC, et al. The worldwide costs of dementia 2015 and comparisons with 2010. Alzheimer’s Dement. 2017;13(1):1–7. doi:10.1016/j.jalz.2016.07.150

13. Schoene D, Wu SMS, Mikolaizak AS, et al. Discriminative ability and predictive validity of the timed up and go test in identifying older people who fall: Systematic review and meta-analysis. J Am Geriatr Soc. 2013;61(2):202–208. doi:10.1111/jgs.12106

14. Jongsiriyanyong S, Limpawattana P. Mild Cognitive Impairment in Clinical Practice: A Review Article. Am J Alzheimers Dis Other Demen. 2018;33(8):500–507. doi:10.1177/1533317518791401

15. Arevalo-Rodriguez I, Smailagic N, Roqué-Figuls M, et al. Mini-Mental State Examination (MMSE) for the early detection of dementia in people with mild cognitive impairment (MCI). Cochrane Database Syst Rev. 2021;2021(7). doi:10.1002/14651858.CD010783.pub3

16. Muir SW, Gopaul K, Montero Odasso MM. The role of cognitive impairment in fall risk among older adults: A systematic review and meta-analysis. Age Ageing. 2012;41(3):299–308. doi:10.1093/ageing/afs012

17. Montero-Odasso M, Verghese J, Beauchet O, Hausdorff JM. Gait and cognition: A complementary approach to understanding brain function and the risk of falling. J Am Geriatr Soc. 2012;60(11):2127–2136. doi:10.1111/j.1532-5415.2012.04209.x

18. Ponce-Bobadilla AV, Schmitt V, Maier CS, Mensing S, Stodtmann S. Practical guide to SHAP analysis: Explaining supervised machine learning model predictions in drug development. Clin Transl Sci. 2024;17(11):1–15. doi:10.1111/cts.70056

19. Hildt E. What Is the Role of Explainability in Medical Artificial Intelligence? A Case-Based Approach. Bioengineering. 2025;12(4). doi:10.3390/bioengineering12040375

20. Patel S, Park H, Bonato P, Chan L, Rodgers M. A review of wearable sensors and systems with application in rehabilitation. J Neuroengineering Rehabil. 2012;9(21):1–17. 10.1186/1743-0003-9-21

21. Godfrey A. Wearables for independent living in older adults: Gait and falls. Maturitas. 2017;100:16–26. doi:10.1016/j.maturitas.2017.03.317

22. Hall JB, Akter S, Rao P, et al. Feasibility of Using a Novel, Multimodal Motor Function Assessment Platform with Machine Learning to Identify Individuals with Mild Cognitive Impairment. Alzheimer Dis Assoc Disord. 2024;38(4):344–350. doi:10.1097/WAD.0000000000000646

23. Makino K, Lee S, Bae S, et al. Simplified Decision-Tree Algorithm to Predict Falls for Community-Dwelling Older Adults. Published online 2021.

24. Reiss A, Stricker D. Confidence-based Multiclass AdaBoost for Physical Activity Monitoring. Published online 2013:13–20. doi:10.1145/2493988.2494325

25. Chen T, Guestrin C. XGBoost: A scalable tree boosting system. Proc ACM SIGKDD Int Conf Knowl Discov Data Min. 2016;13–17-Augu:785-794. doi:10.1145/2939672.2939785

26. Shickel B, Tighe PJ, Bihorac A, Rashidi P. Deep EHR: A Survey of Recent Advances in Deep Learning Techniques for Electronic Health Record (EHR) Analysis. IEEE J Biomed Heal Informatics. 2018;22(5):1589–1604. doi:10.1109/JBHI.2017.2767063

27. Scott M. Lundberg, Su-In Lee. A Unified Approach to Interpreting Model Predictions. Nips. 2017;16(3):426–430.

28. S. Rasoul Safavian DL. A survey of decision tree classifier methodology - Systems, Man and Cybernetics, IEEE Transactions on. 1991;21(3):660–674. doi:10.9790/5933-0612104113

29. Hellmers S, Krey E, Gashi A, et al. Comparison of machine learning approaches for near-fall-detection with motion sensors. Front Digit Heal. 2023;5(July):1–10. doi:10.3389/fdgth.2023.1223845

30. Noh B, Youm C, Goh E, et al. XGBoost based machine learning approach to predict the risk of fall in older adults using gait outcomes. Sci Rep. 2021;11(1):1–9. doi:10.1038/s41598-021-91797-w

31. Subasi A, Dammas DH, Alghamdi RD, et al. Sensor based human activity recognition using adaboost ensemble classifier. Procedia Comput Sci. 2018;140:104–111. doi:10.1016/j.procs.2018.10.298

32. Lockhart TE, Soangra R, Yoon H, et al. Prediction of fall risk among community-dwelling older adults using a wearable system. Sci Rep. 2021;11(1):1–14. doi:10.1038/s41598-021-00458-5

33. Safavian SR, Landgrebe D. A Survey of Decision Tree Classifier Methodology. IEEE Trans Syst Man Cybern. 1991;21(3):660–674. doi:10.1109/21.97458

34. Selection R. A Machine Learning Multi-Class Approach for Fall Detection Rates Selection †. Published online 2021. doi:10.1109/ICAIIC48513.2020.9065205.Citation

35. Özdemir AT, Barshan B. Detecting Falls with Wearable SensorsUsing Machine Learning Techniques. Published online 2014:10691–10708. doi:10.3390/s140610691

36. Subramaniam S, Faisal AI, Deen MJ. Wearable Sensor Systems for Fall Risk Assessment : A Review. 2022;4(July). doi:10.3389/fdgth.2022.921506

37. Lazarou E, Exarchos TP. Predicting stress levels using physiological data : Real-time stress prediction models utilizing wearable devices. 2024;11(December 2023):76–102. doi:10.3934/Neuroscience.2024006

38. Tripathy RK, Paternina MA, Antonio J, Serna DO. Editorial : Machine Learning and Deep Learning for Physiological Signal Analysis. 2022;13(April):3389–3390. doi:10.3389/fphys.2018.00722

39. Amann J, Blasimme A, Vayena E, Frey D, Madai VI. Explainability for artificial intelligence in healthcare: a multidisciplinary perspective. BMC Med Inform Decis Mak. 2020;20(1):1–9. doi:10.1186/s12911-020-01332-6

40. Grote T, Berens P. On the ethics of algorithmic decision-making in healthcare. J Med Ethics. 2020;46(3):205–211. doi:10.1136/medethics-2019-105586

41. Nasreddine ZS, Phillips NA, Bédirian V, et al. The Montreal Cognitive Assessment, MoCA: A brief screening tool for mild cognitive impairment. J Am Geriatr Soc. 2005;53(4):695–699. doi:10.1111/j.1532-5415.2005.53221.x

42. Kakara R, Bergen G, Burns E, Stevens M. Nonfatal and Fatal Falls Among Adults Aged ≥65 Years — United States, 2020–2021. MMWR Morb Mortal Wkly Rep. 2023;72(35):938–943. doi:10.15585/mmwr.mm7235a1

43. Gobbens RJJ, van Assen MALM, Luijkx KG, Wijnen-Sponselee MT, Schols JMGA. The tilburg frailty indicator: Psychometric properties. J Am Med Dir Assoc. 2010;11(5):344–355. doi:10.1016/j.jamda.2009.11.003

44. Guess TM, Bliss R, Hall JB, Kiselica AM. Gait & Posture Comparison of Azure Kinect overground gait spatiotemporal parameters to marker based optical motion capture. Gait Posture. 2022;96(September 2021):130–136. doi:10.1016/j.gaitpost.2022.05.021

45. Thomas J, Hall JB, Bliss R, Guess TM. Comparison of Azure Kinect and optical retroreflective motion capture for kinematic and spatiotemporal evaluation of the sit-to-stand test. Gait Posture. 2022;94(November 2021):153–159. doi:10.1016/j.gaitpost.2022.03.011

46 . Thomas JM, Hall JB, Bliss R, et al. A machine learning approach to concussive group classification using discrete outcome measures from a low-cost movement-based assessment system. 2025;144(July 2024).

47. Pyle D, Editor S, Cerra DD. Data Preparation for Data Mining.; 1999.

48. Pedregosa F, Weiss R, Brucher M. Scikit-learn : Machine Learning in Python. 2011;12:2825–2830.

49. Kang H. The prevention and handling of the missing data. 2013;64(5):402–406.

50. Chawla N V, Bowyer KW, Hall LO, Kegelmeyer WP. SMOTE : Synthetic Minority Over-sampling Technique. 2002;16:321–357.

51. Li Y, Yang Y, Song P, Duan L, Ren R. An improved SMOTE algorithm for enhanced imbalanced data classification by expanding sample generation space. Published online 2025:1–21.

52. Shipe ME, Deppen SA, Farjah F, Grogan EL. Developing prediction models for clinical use using logistic regression: An overview. J Thorac Dis. 2019;11(Suppl 4):S574–S584. doi:10.21037/jtd.2019.01.25

53. Javeed A, Dallora AL, Berglund JS, Anderberg P. An Intelligent Learning System for Unbiased Prediction of Dementia Based on Autoencoder and Adaboost Ensemble Learning. Life. 2022;12(7). doi:10.3390/life12071097

54. Ramírez F, Gutiérrez M. Dual-Task Gait as a Predictive Tool for Cognitive Impairment in Older Adults: A Systematic Review. Front Aging Neurosci. 2021;13(December):1–13. doi:10.3389/fnagi.2021.769462

55. Zhou J, Habtemariam D, Iloputaife I, Lipsitz LA, Manor B. The complexity of standing postural sway associates with future falls in community-dwelling older adults: The MOBILIZE Boston study. Sci Rep. 2017;7(1):1–8. doi:10.1038/s41598-017-03422-4

56. Baltasar-Fernandez I, Alcazar J, Mañas A, et al. Relative sit-to-stand power cut-off points and their association with negatives outcomes in older adults. Sci Rep. 2021;11(1):1–10. doi:10.1038/s41598-021-98871-3

57. Akter S, Liu Z, Simoes EJ, Rao P. Machine Learning for Early Prediction of Alzheimer’s Disease and Related Dementias Using Electronic Health Record (EHR) Data. En: American Medical Informatics Association (AMIA) Informatics Summit, March 2025, Pittsburgh, PA (Poster).; 2025. doi:https://amia.secure-platform.com/summit/gallery/rounds/82009/details/14794

58. Akter S, Liu Z, Simoes EJ, Rao P. Using machine learning and electronic health record (EHR) data for the early prediction of Alzheimer’s Disease and Related Dementias. J Prev Alzheimer’s Dis. 2025;12(April):100169. doi:10.1016/j.tjpad.2025.100169

