## Supplementary Material for "Explainable Machine Learning for Early Detection of Mild Cognitive Impairment, Fall Risk, and Frailty Using Sensor-Based Motor Function Data" (from another version of this preprint: version 1, 2025-12-26)

### Supplementary Materials

#### Supplementary Methods: Feature Engineering and Model Inputs

This section provides a complete description of the features used as inputs to the machine learning models. While the Data Collection section describes the assessment tasks and sensor modalities, the features listed here represent the final engineered variables included in model development following preprocessing and quality control. Features are grouped by task and experimental condition (single-task/ST and dual-task/DT) for clarity.

##### Supplementary Table S1. Gait Features Used for Model Development

| Feature category | Feature description | Sensor modality | Condition |
|---|---|---|---|
| Spatiotemporal | Stride length | Depth camera | ST, DT |
| Spatiotemporal | Stride time | Depth camera | ST, DT |
| Spatiotemporal | Step width | Depth camera | ST, DT |
| Spatiotemporal | Step length | Depth camera | ST, DT |

##### Supplementary Table S2. Sit-to-Stand (STS) Features Used for Model Development

| Feature category | Feature description | Sensor modality | Condition |
|---|---|---|---|
| Spatiotemporal | Time-to-completion (3 repetitions) | Depth camera | ST, DT |
| Spatiotemporal | Time-to-completion (5 repetitions) | Depth camera | ST, DT |
| Kinematic | Average knee angular velocity (sit phase) | Depth camera | ST, DT |
| Kinematic | Average knee angular velocity (stand phase) | Depth camera | ST, DT |

##### Supplementary Table S3. Static Balance Features Used for Model Development

| Feature category | Feature description | Sensor modality | Condition |
|---|---|---|---|
| Postural sway | CoP sway area (95%) | Force plate | ST, DT |
| Postural sway | Mean CoP velocity | Force plate | ST, DT |
| Directional sway | CoP velocity (AP) | Force plate | ST, DT |
| Directional sway | CoP velocity (ML) | Force plate | ST, DT |
| Postural sway | CoM sway (AP, ML) | Depth camera | ST, DT |

##### Supplementary Table S4. Step-Down Features Used for Model Development

| Feature category | Feature description | Sensor modality |
|---|---|---|
| Temporal | Leading limb time | Depth camera |
| Temporal | Trailing limb time | Depth camera |
| Stability | Medial–lateral CoM sway | Depth camera |

##### Supplementary Table S5. Reaction Time and Grip Strength Features Used for Model Development

| Feature category | Feature description | Sensor modality |
|---|---|---|
| Reaction time | Mean response time | Interface board |
| Accuracy | Response accuracy | Interface board |
| Strength | Mean bilateral grip strength | Dynamometer |
| Symmetry | Grip strength asymmetry index | Dynamometer |

All features listed above were standardized prior to model training and were used consistently across single-outcome and unified multi-label models unless otherwise specified.

Supplementary Table S6: Model configurations and hyperparameters for all ML models (MCI, Fall Risk, Frailty, and Unified Multilabel Framework).

| **Mild Cognitive Impairment (MCI) Models** | | |
|---|---|---|
| **Model** | **Algorithm** | **Best Hyperparameters** |
| Baseline | LR | C = 10; class_weight = None; fit_intercept = True; l1_ratio = 0.0; max_iter = 2000; penalty = 'l1'; solver = liblinear. |
| Optimized (All features) | XGBoost | colsample_bytree = 0.4; learning_rate = 0.4; max_depth = 4; n_estimators = 150; subsample = 0. |
| Best | XGBoost | colsample_bytree = 0.4; learning_rate = 0.4; max_depth = 4; n_estimators = 185; subsample = 0.2. |
| **Fall-Risk Models** | | |
| **Model** | **Algorithm** | **Best Hyperparameters** |
| Baseline | LR | C = 0.001; class_weight = None; l1_ratio = 0; max_iter = 500; penalty = 'l2'; solver = liblinear. |
| Optimized (All features) | XGBoost | colsample_bytree = 0.8; gamma = 0; learning_rate = 0.05; max_bin = 256; max_depth = 3; min_child_weight = 1; n_estimators = 1200; subsample = 1.0 |
| Best | XGBoost | colsample_bytree = 0.8; gamma = 0; learning_rate = 0.05; max_bin = 256; max_depth = 3; min_child_weight = 1; n_estimators = 1200; subsample = 1.0; random_state = 60. |
| **Frailty Models** | | |
| **Model** | **Algorithm** | **Best Hyperparameters** |
| Baseline | LR | C = 0.1; class_weight = balanced; l1_ratio = 0; max_iter = 500; penalty = l1; solver = liblinear |
| Optimized (All features) | XGBoost | colsample_bytree: 0.8, gamma: 0, learning_rate: 0.05, max_bin: 256, max_depth: 2, min_child_weight: 2, n_estimators: 800, subsample: 0.8 |
| Best | Decision Tree | ccp_alpha = 0.0; class_weight = None; criterion = gini; max_depth = 3; max_features = log2; min_samples_leaf = 1; min_samples_split = 2; splitter = random |
| **Unified Multilabel Model (MCI + Fall-Risk + Frailty)** | | |
| **Model** | **Algorithm** | **Best Hyperparameters** |
| Optimized | XGBoost | colsample_bytree = 0.8599; learning_rate = 0.0258; max_bin = 501; max_depth = 3; min_child_weight = 5; n_estimators = 1101; reg_lambda = 1.2435; scale_pos_weight = 1.7930; subsample = 0.8503. |
| Alternative | Decision Tree | class_weight = balanced; criterion = entropy; max_depth = 4; max_features = log2; min_samples_leaf = 2; min_samples_split = 10. |
| Alternative | AdaBoost | estimator__max_depth = 1; estimator__min_samples_leaf = 3; learning_rate = 0.2294; n_estimators = 136. |

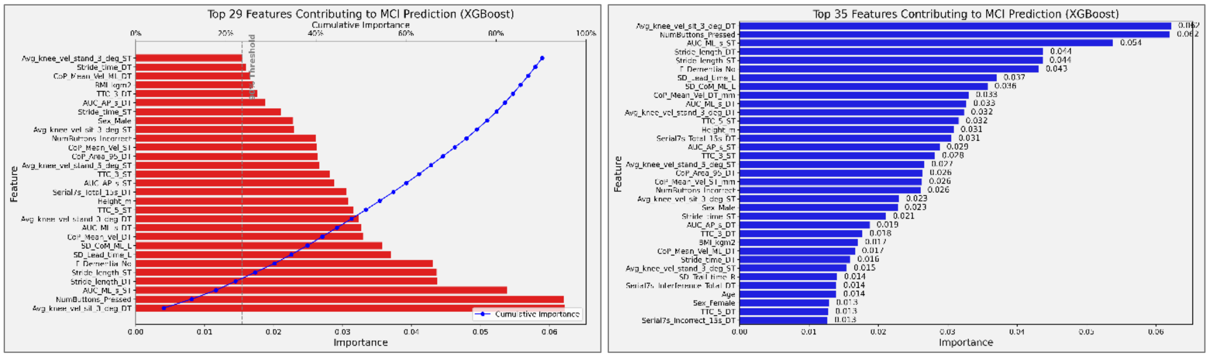


Supplementary Figure S7: Cumulative feature importance of MCI prediction.

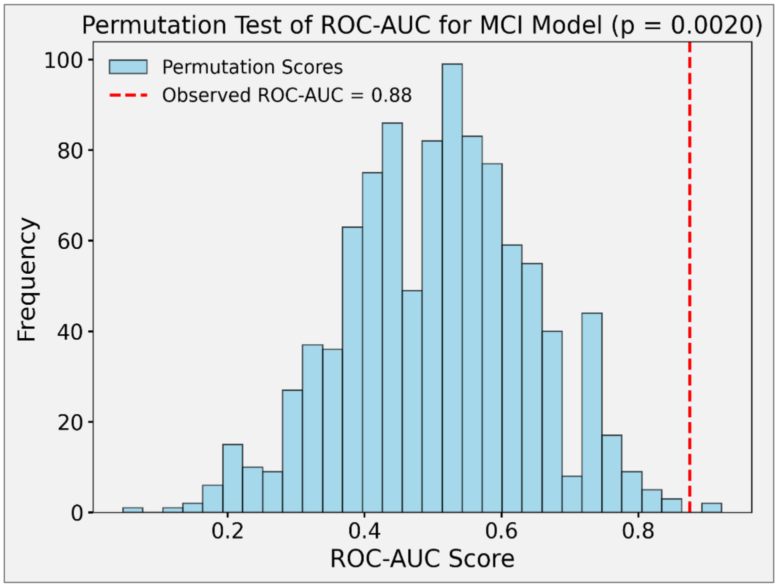


Supplementary Figure S8: Permutation Test for MCI Model Significance (ROC-AUC).

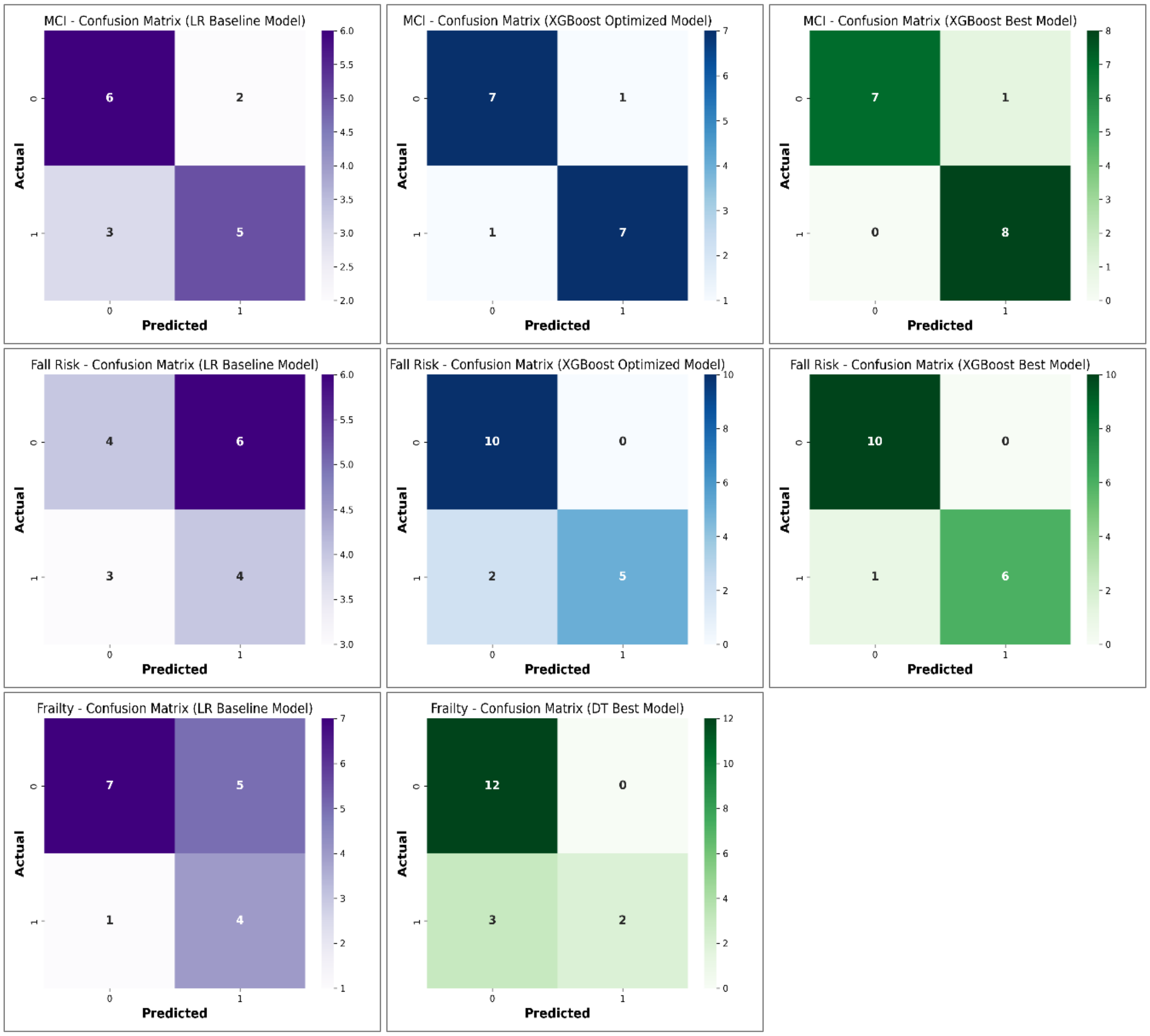


Supplementary Figure S9: Confusion matrices illustrating the classification performance of baseline, optimized (all features), and best-performing models for MCI, fall risk, and frailty prediction.

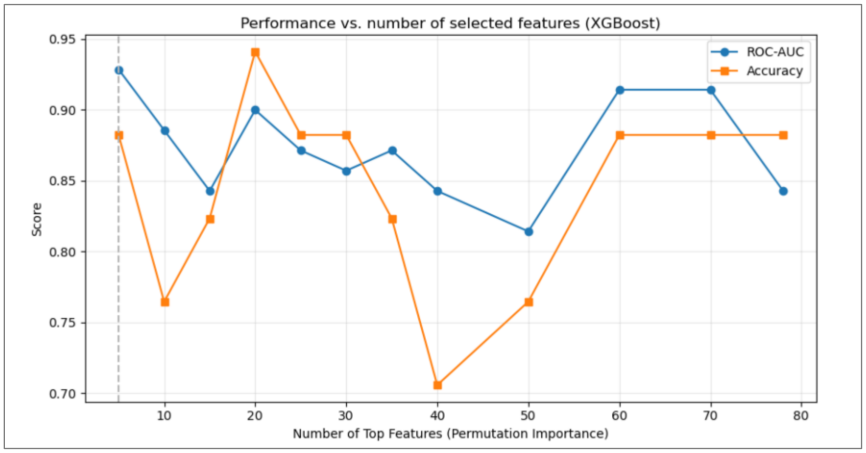


Supplementary Figure S10: Fall Risk Model Performance Across Different Numbers of Top Features (Permutation Importance).

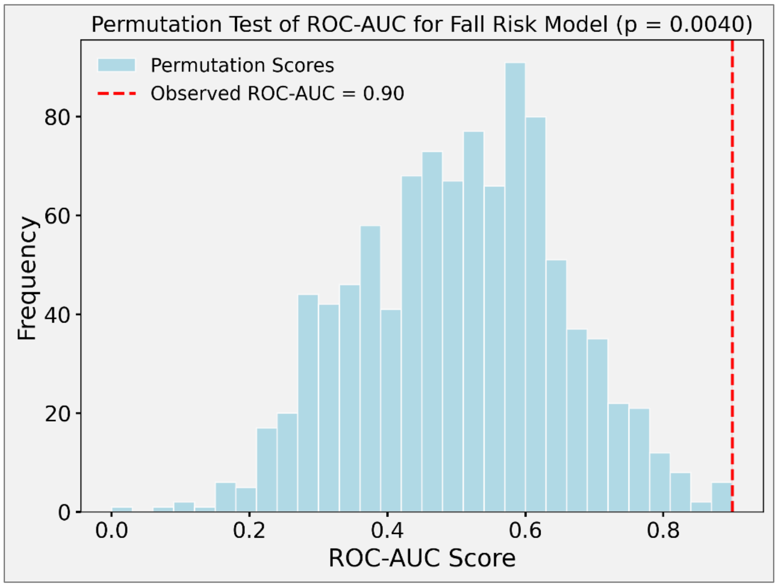


Supplementary Figure S11: Permutation Test for Fall Risk Model Significance (ROC-AUC).

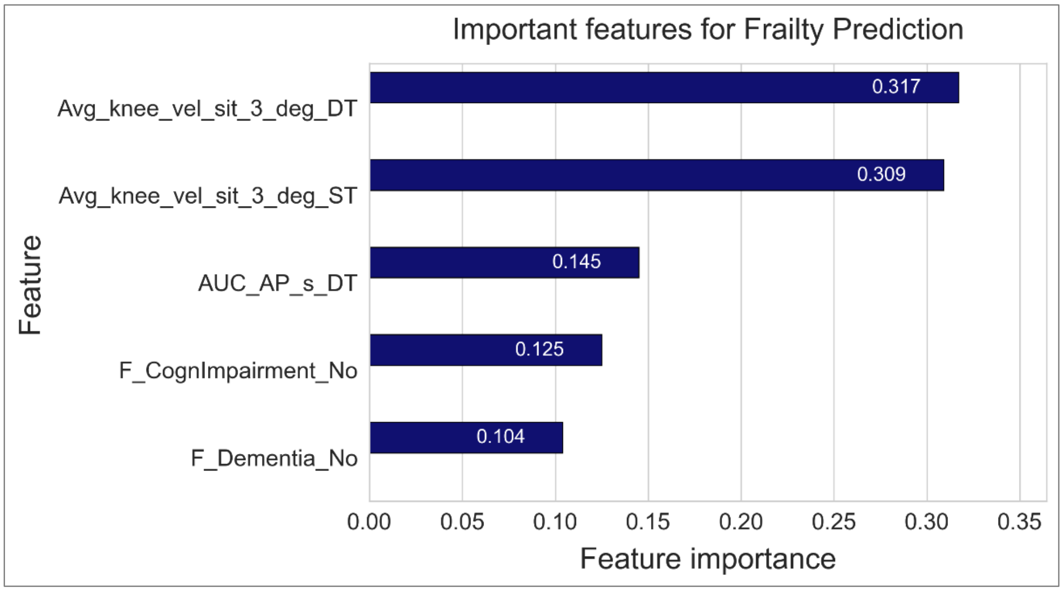


Supplementary Figure S12: Feature importance analysis of Frailty Model

Supplementary Table S13: Detailed performance of unified multilabel models across classifiers and outcomes (MCI, Frailty, and Fall Risk).

| **Model** | **Metrics** | | | | |
|---|---|---|---|---|---|
| **Decision Tree** | **Accuracy** | **AUC** | **Recall** | **Specificity** | **F1 Score** |
| MCI | 0.88 | 0.92 | 0.71 | 1.00 | 0.86 |
| Fall Risk | 0.55 | 0.63 | 0.29 | 0.67 | 0.35 |
| Frailty | 0.62 | 0.57 | 0.40 | 0.73 | 0.40 |
| **XGBoost** | **Accuracy** | **AUC** | **Recall** | **Specificity** | **F1 Score** |
| MCI | 0.81 | 0.71 | 0.71 | 0.89 | 0.77 |
| Fall Risk | 0.69 | 0.67 | 0.71 | 0.68 | 0.67 |
| Frailty | 0.69 | 0.51 | 0.20 | 0.91 | 0.70 |
| **AdaBoost** | **Accuracy** | **AUC** | **Recall** | **Specificity** | **F1 Score** |
| MCI | 0.81 | 0.79 | 0.71 | 0.89 | 0.77 |
| Fall Risk | 0.63 | 0.54 | 0.43 | 0.78 | 0.52 |
| Frailty | 0.69 | 0.62 | 0.20 | 0.91 | 0.35 |

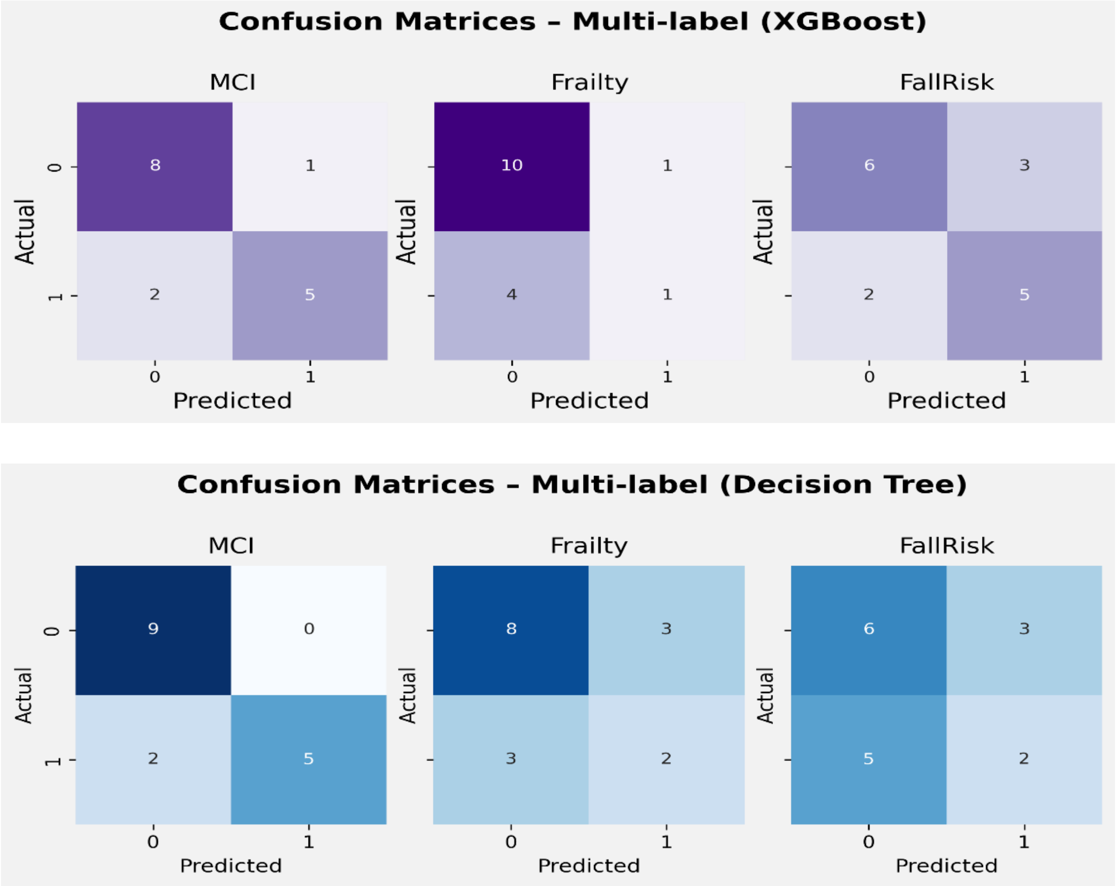


Supplementary Figure S14: Confusion matrices of unified multilabel models for MCI, Frailty, and Fall Risk using XGBoost (top) and Decision Tree (bottom) classifiers.

### Feature Abbreviations and Definitions

#### Demographic Variables

Age – Participant age (years)

Height_m – Height (meters)

Weight_kg – Body weight (kilograms)

BMI_kgm2 – Body mass index (kg/m²)

Sex – Biological sex

Race – Self-reported race

#### Clinical and Health History Indicators

F_Alzheimers – Family history of Alzheimer's disease

F_Dementia – Family history of dementia

F_Parkinsons – Family history of Parkinson's disease

F_Stroke – Family history of stroke

F_CognImpairment – Family history of cognitive impairment

F_Schizophrenia – Family history of schizophrenia

F_Bipolar – Family history of bipolar disorder

F_Alcholism – Family history of alcoholism

#### Gait Features

Stride_length_avg_nll_ST – Mean stride length during single-task walking

Stride_length_avg_nll_DT – Mean stride length during dual-task walking

Stride_time_avg_s_ST – Mean stride time during single-task walking

Stride_time_avg_s_DT – Mean stride time during dual-task walking

#### Step Down (SD) Features

SD_Lead_time_L_s – Mean step-down time of the leading limb (left)

SD_Lead_time_R_s – Mean step-down time of the leading limb (right)

SD_Trail_time_L_s – Mean step-down time of the trailing limb (left)

SD_Trail_time_R_s – Mean step-down time of the trailing limb (right)

SD_CoM_ML_L_mm – Mean step-down mediolateral center-of-mass displacement (left)

SD_CoM_ML_R_mm – Mean step-down mediolateral center-of-mass displacement (right)

**Static Balance Features**

CoP_Area_95_sqmm_ST – 95% confidence ellipse area of center-of-pressure sway during single-task balance assessment

CoP_Area_95_sqmm_DT – 95% confidence ellipse area of center-of-pressure sway during dual-task balance assessment

CoP_Mean_Vel_mm_s_ST – Mean velocity of center-of-pressure displacement during single-task balance assessment

CoP_Mean_Vel_mm_s_DT – Mean velocity of center-of-pressure displacement during dual-task balance assessment

CoP_Mean_Vel_ML_mm_s_ST – Mean mediolateral center-of-pressure velocity during single-task balance assessment

CoP_Mean_Vel_ML_mm_s_DT – Mean mediolateral center-of-pressure velocity during dual-task balance assessment

CoP_Mean_Vel_AP_mm_s_ST – Mean anteroposterior center-of-pressure velocity during single-task balance assessment

CoP_Mean_Vel_AP_mm_s_DT – Mean anteroposterior center-of-pressure velocity during dual-task balance assessment

AUC_AP_mm_s_ST – Area under the curve of the anteroposterior balance stability signal during single-task condition

AUC_AP_mm_s_DT – Area under the curve of the anteroposterior balance stability signal during dual-task condition

AUC_ML_mm_s_ST – Area under the curve of the mediolateral balance stability signal during single-task condition

AUC_ML_mm_s_DT – Area under the curve of the mediolateral balance stability signal during dual-task condition

Serial7s_Total_15s_DT – Total number of Serial 7s responses during the dual-task balance condition (15 seconds)

Serial7s_Incorrect_15s_DT – Number of incorrect Serial 7s responses during the dual-task balance condition (15 seconds)

Serial7s_Interference_Total_DT – Dual-task interference score based on total Serial 7s responses during balance assessment

Serial7s_Interference_Correct_DT – Dual-task interference score based on correct Serial 7s responses during balance assessment

**Sit-to-Stand Features**

Avg_knee_vel_stand_3_deg_s_ST – Average knee angular velocity during the stand phase (3° threshold) under single-task condition
Avg_knee_vel_stand_3_deg_s_DT – Average knee angular velocity during the stand phase (3° threshold) under dual-task condition
Avg_knee_vel_sit_3_deg_s_ST – Average knee angular velocity during the sit phase (3° threshold) under single-task condition
Avg_knee_vel_sit_3_deg_s_DT – Average knee angular velocity during the sit phase (3° threshold) under dual-task condition
Avg_knee_vel_stand_5_deg_s_ST – Average knee angular velocity during the stand phase (5° threshold) under single-task condition
Avg_knee_vel_stand_5_deg_s_DT – Average knee angular velocity during the stand phase (5° threshold) under dual-task condition
Avg_knee_vel_sit_5_deg_s_ST – Average knee angular velocity during the sit phase (5° threshold) under single-task condition
Avg_knee_vel_sit_5_deg_s_DT – Average knee angular velocity during the sit phase (5° threshold) under dual-task condition
TTC_3_s_ST – Time-to-contact during sit-to-stand computed using a 3-second window under single-task condition
TTC_3_s_DT – Time-to-contact during sit-to-stand computed using a 3-second window under dual-task condition
TTC_5_s_ST – Time-to-contact during sit-to-stand computed using a 5-second window under single-task condition
TTC_5_s_DT – Time-to-contact during sit-to-stand computed using a 5-second window under dual-task condition

**Reaction Time Features**

Reaction_Time_ms – Mean reaction time (milliseconds)
NumButtons_Pressed – Total number of button presses during the reaction task
NumButtons_Incorrect – Number of incorrect button presses during the reaction task
